# Estradiol, des-acylated, and total ghrelin levels might be associated with epithelial ovarian cancer in postmenopausal women

**DOI:** 10.1101/2020.04.26.20080440

**Authors:** Saba Fooladi, Hamed Akbari, Moslem Abolhassani, Erfan Sadeghi, Hossein Fallah

## Abstract

**Background:** The present study aimed to investigate the association between estradiol, acylated, des-acylated, total ghrelin levels along with pathological parameters and epithelial ovarian cancer (EOC) odds in postmenopausal women.

**Methods:** A case-control study was carried out on 45 EOC patients and 33 age-matched postmenopausal women as the control group. Plasma levels of estradiol, acylated, des-acylated, and total ghrelin were measured by ELISA method.

**Results:** Estradiol’s plasma levels were significantly higher in EOC patients compared to control women (*P* < 0.001). Although acylated, des-acylated, and total ghrelin levels were not associated with EOC in logistic regression models, estradiol levels were significantly related to the increase in EOC odds (OR: 1.083, 95% CI: 1.037-1.13, *P* < 0.001). However, estradiol levels in the two first quartiles (Q_1_, Q_2_) were associated with decreased odds of EOC (OR: 0.011, 95% CI: 0.001-0.118, *P* < 0.001, and OR: 0.030, 95% CI: 0.003-0.284, *P* = 0.002, respectively). For those patients in the third quartile of plasma des-acylated and total ghrelin compared to those in the highest (Q4), the multivariate odds ratios of EOC were respectively 0.192 and 0.25.

**Conclusions:** In conclusion, higher concentrations of des-acylated and total ghrelin might be associated with the decreased EOC odds. Furthermore, the findings suggest that high levels of estradiol might be a potential odds factor in EOC, however, lower estradiol levels may have a protective effect on EOC development.

## INTRODUCTION

Ovarian cancer (OC) accounts for the most clinical challenges and mortality rate among all gynecological cancers and epithelial ovarian cancer (EOC) comprises 90% of all OCs (1, 2). Due to remaining asymptomatic until metastasis, OC is the most current ovarian malignancy diagnosed in advanced stages in more than two-thirds of patients (3). Subsequently, studies demonstrated that the primary stage diagnosis can reduce the mortality rate to 50% and increase the survival rate up to 95% (4–6). Numerous factors interfere with OC risk including fertility history, reproduction period (menstrual interval), oral contraceptive pill (OCP) consumption, and breastfeeding which are associated with OC risk through hormone-dependent mechanisms (7).

Estrogen and progesterone are feminine hormones which play an important role in the fertility process. The presence of estrogen in females increases cancer risk, especially in hormone target tissues such as ovary, endometrium, and breast (8). Meanwhile, epidemiologic and *in-vitro* studies confirmed the detrimental role of estrogen in OC through the carcinogenic effects of estrogen accumulation during continual ovulation cycles (9). It is also well known that estrogen not only increases cell proliferation and ovarian growth but also converts normal ovary cells to malignant ones, which reflects the estrogen effect on OC development (9). Several studies demonstrated that estrogen receptor − α positive (ER^+^) malignant cells proliferate in response to increase in estrogen (10, 11). Precisely, many studies assert that ER-β prevails in intact and benign tumors, while ER-α is the major type of ER in malignant cells, which increase the proportion of ER-α to ER-β in OC (11, 12).

Ghrelin is a peptide hormone containing 28 amino acids produced and secreted along with the digestive system (13). There are two common forms of ghrelin in blood including acylated and des-acylated forms, which consist of 70%-80% of total ghrelin. The acylated peptide specifically releases growth hormone both *in-vivo* and *in-vitro* and is essential for its activity (14). Ghrelin and its receptor are expressed in some cancers and cancerous cell lines, such as stomach, intestine, kidney, esophagus, breast, prostate, and OC. On the other hand, studies indicated the effects of mentioned hormones on cancer development through cell proliferation, apoptosis, tissue invasion, and metastasis (15). Growth hormone secretagogue receptor 1a and 1b (GSHR-1a and GHSR-1b) are the major types of ghrelin receptors, and studies represent ghrelin and GSHR-1a relation in OC, however, studies in this subject are limited and sometimes contradictory.

An ample body of evidence has introduced ghrelin as an apoptotic and autophagocytosis factor in human ovarian cancerous cells and has shown the role of ovarian epithelium and ovarian associated tumors in systemic or local ghrelin generation due to the expression of GHSR-1a in ovaries (16–19). Although some studies indicated ghrelin role in the incidence of some cancers such as stomach and esophagus as well as the importance of measuring ghrelin as an early biomarker in the diagnosis of these cancers, published articles in this field placed an emphasis on studying molecular aspects of ghrelin and ovarian malignant tumors. Moreover, there is little and inconsistent data regarding the association of estrogen, ghrelin and pathological findings in postmenopausal EOC patients. Therefore, the present study aimed to investigate the association between estradiol, acylated, des-acylated, and total ghrelin levels along with pathological parameters, on the one hand, and EOC odds in postmenopausal women, on the other.

## METHODS

### Participants

This case-control study was conducted on 45 postmenopausal female volunteers, suffering from EOC, who referred to the Iran National Tumor Bank, which is funded by the Cancer Institute of Tehran University of Medical Sciences for Cancer Research, Tehran, Iran. All EOC participants were newly diagnosed and EOC tumors were confirmed through histopathological studies. Thirty-three cancer-free and healthy age-matched postmenopausal women with no previous history of the disease as well as no consumption of any medication were included as the control group. Of 47 healthy menopause women who were invited to participate in the study, 33 (70%) agreed to participate in the study and were included as the control group. The case group inclusion criteria were as follows: post-menopausal status, proper operation of body major organs, the absence of any other malignant tumors in the body except ovarian one, the absence of starting any treatment, including surgery, hormonal therapy, and chemotherapy. Moreover, exclusion criteria included participants’ dissatisfaction, neoplastic, metabolic and cardiovascular disorders or other concurrent medical illness such as renal disease, acute or chronic infection, and hepatic disorders. Almost all the women were also non-smokers (except one woman). Written informed consents, as an important requirement for providing participants with the advice and the details on the proposed trial, were obtained from all participants. The study conforms to the *Declaration of Helsinki* regarding research involving human subjects and is approved by the Ethics Committee of Kerman University of Medical Sciences, Kerman, Iran (IR.KMU.REC.1394.193).

### Samples and data collection

Demographic data were collected using a questionnaire and 5 ml blood sample was drawn in K_2_EDTA sample tubes from each participant. Blood samples were immediately centrifuged at 4°C in order to separate plasma. All the plasma samples were stored at -70°C until analysis. The fasting blood sugar (FBS), urea, creatinine (Cr), and blood lipids such as total cholesterol (TC), triglycerides (TG), high-density lipoprotein cholesterol (HDL-c), and low-density lipoprotein cholesterol (LDL-c) were all determined by standard kits (Pars Azmoon, Tehran, Iran) by an autoanalyzer (Hitachi 902 autoanalyzer; Roche Diagnostics, Mannheim, Germany) in a standard laboratory setting.

### Hormonal assays

The basal serum levels of acylated ghrelin, des-acylated ghrelin, and estradiol were measured using the enzyme immunometric assay (EIA) kit (Biovendor, Karasek, Brno, Czech Republic), with a lower limit detection of 6 pg/ml. Moreover, plasma estradiol levels were determined by EIA kit (Accubind ELISA microwells, Monobind Inc, Lake Forest, California, USA) with a lower limit of detection of 8.2 pg/ml. Experiments were performed according to the manufacturer’s protocol.

### Statistical analysis

Continues and categorical variables were mentioned in terms of mean ± SD (standard deviation) and number (percent), respectively. The logistic regression model was used to investigate the association between the four main variables and EOC odds. Variables were introduced continuously and qualitatively (classified according to first to third quartiles) into the model. Then, the estimated odds ratio (OR) was calculated in two crude and adjusted modes (based on the age variable). To examine the linear relationship between these variables and other blood and pathological parameters, Spearman’s correlation coefficient was used. Moreover, Mann-Whitney and Kruskal-Wallis tests were employed to examine the relationship between the main variables and tumor stages as well as tumor grades. The statistical analyses were carried out using SPSS software version 24.0 for Windows (IBM/SPSS Inc., New York, USA). *P*-values less than 0.05 were considered statistically significant.

## RESULTS

### Demographic analysis

In the present study, we measured the estradiol, acylated, and des-acylated ghrelin levels of 45 EOC and 33 healthy women. Comparison of demographic, clinical, and pathological variables between EOC and control women are summarized in Table 1. The median age of patients was 60 years and the case and control groups did not show a significant statistical difference in terms of age and menopausal status (*P* > 0.05 for both comparisons). Forty percent of EOC patients were in early cancer stage, and 28% of them were in advanced one. Furthermore, three histological EOC grades were determined among patients, suggesting that the distribution of grade I, II, and III was 22.2%, 22.2%, and 26.7%, respectively. The patients’ tumor size was divided into three major categories including: T_1_, T_2_, and T_3_. T_1_ stands for the tumors under 2 cm, T_2_ for tumors between 2 and 5 cm, and T_3_ for tumors beyond 5 cm. The distribution of T_1_, T_2_, and T_3_ was 33.3%, 13.3%, and 22.2%, respectively.

**Table 1.** Comparison of demographic, clinical and pathological parameters between EOC and control groups.

| Variables |  | Control<br>n = 33 | EOC<br>n = 45 | P-value |
| --- | --- | --- | --- | --- |
| Age of patients<br>(year) | < 60 | 23 (70) | 21 (47) | 0.1 |
|  | ≥ 60 | 10 (30) | 24 (53) |  |
| Histological<br>grade | I | ----- | 10 (22.2) | ----- |
|  | II | ----- | 10 (22.2) |  |
|  | III | ----- | 12 (26.7) |  |
| Stage of disease | Early (stage I-II) | ----- | 18 (40) | ----- |
|  | Advanced (stage III –IV) | ----- | 13 (28) |  |
| Tumor size (cm) | T1 | ----- | 15 (33.3) | ----- |
|  | T2 | ----- | 6 (13.3) |  |
|  | T3 | ----- | 10 (22.2) |  |
| Serum<br>hemoglobin<br>level (12 g/dL) | Normal | 25 (85) | 27 (60) | 0.2 |
|  | Low | 5 (15) | 12 (26.7) |  |
| Serum TC level<br>(200 mg/dL) | Normal | 29 (88) | 44 (100) | 0.1 |
|  | High | 4 (12) | 1 (0) |  |
| **Serum TG level**<br>**(200 mg/dL)** | Normal | 29 (88) | 43 (96) | 0.4 |
|  | High | 4 (12) | 2 (4) |  |
| **Serum LDL-c**<br>**level (130**<br>**mg/dL)** | Normal | 30 (91) | 45 (100) | 0.1 |
|  | High | 3 (9) | 0 (0) |  |
| **Serum HDL-c**<br>**level (50 mg/dL)** | Normal | 26 (79) | 42 (100) | 0.1 |
|  | High | 7 (21) | 3 (0) |  |
| **Previous cancer** | Yes | 0 (0) | 2 (4.5) | <0.001 |
|  | No | 33 (100) | 42 (93.3) |  |
| **Smoking status** | Smoker | 0 (0) | 1 (2) | >0.999 |
|  | Non-smoker | 33 (100) | 44 (98) |  |
| **Family history**<br>**of cancer** | Yes | 1 (3) | 15 (33) | >0.999 |
|  | No | 32 (97) | 30 (67) |  |
| **Alcohol intake** | Yes | 0 (0) | 1 (2) | >0.999 |
|  | No | 33 (100) | 44 (98) |  |

EOC, epithelial ovarian cancer; TC, total cholesterol; TG, triglycerides; LDL-c, low-density lipoprotein cholesterol; HDL-c, high-density lipoprotein cholesterol. Values are presented as number (%).

### Ghrelin and estradiol concentrations

No significant differences were detected in terms of acylated (8.64 ± 3.60 and 9.35 ± 4.30, for patients and controls, respectively), des-acylated ghrelin (118.38 ± 63.75 and 105.53 ± 26.41, for patients and controls, respectively), and total ghrelin (127.03 ± 64.58 and 110.27 ± 34.26, for patients and controls, respectively) mean levels between the EOC and control groups (*P* = 0.407 and *P* =0.905, respectively). However, estradiol showed a significant higher mean concentration in case (55.67 ± 23.03) group compared to control ones (32.18 ± 23.31) (*P* < 0.001) (Fig. 1).

**Fig. 1.**
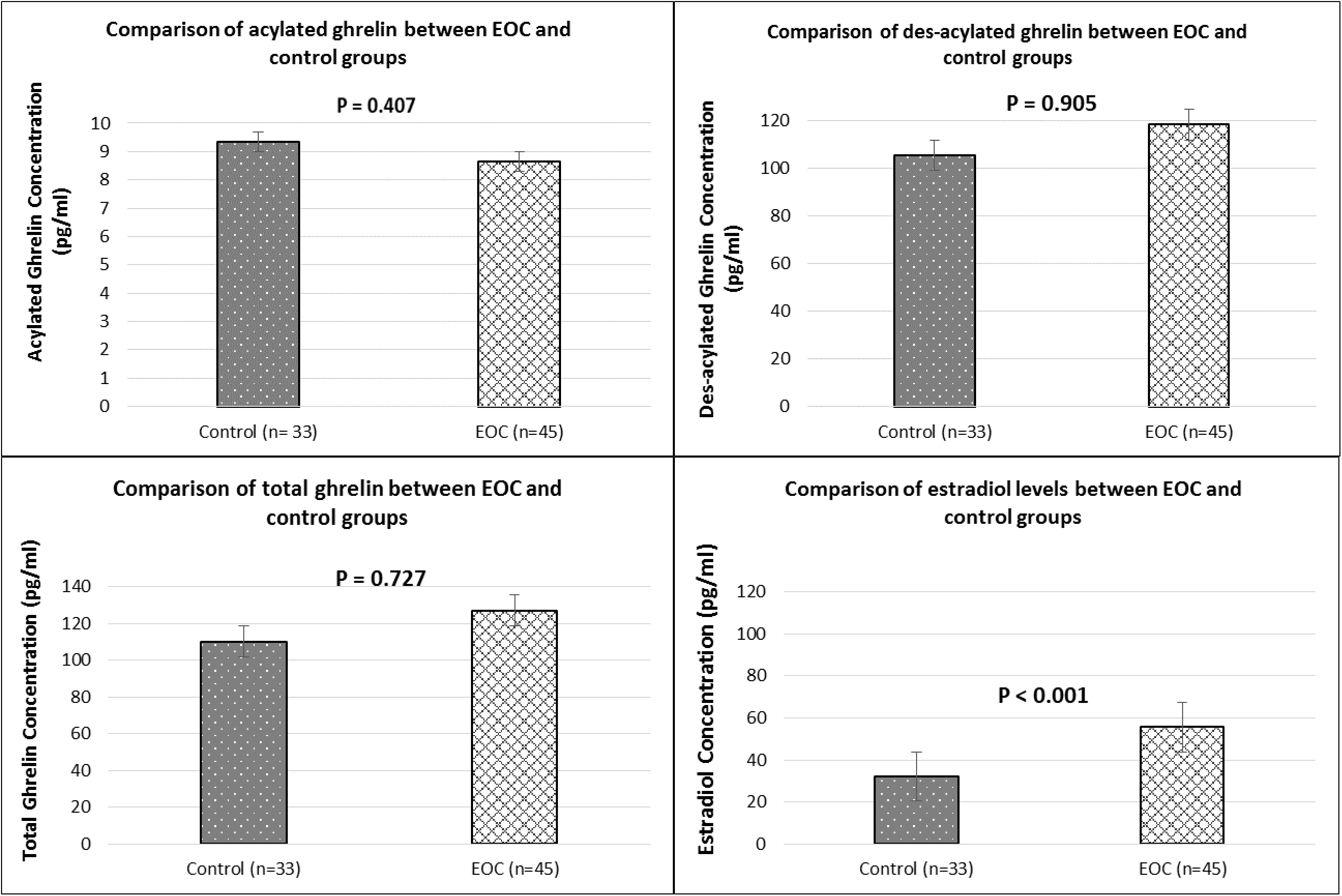
Comparison of acylated ghrelin, des-acylated ghrelin, total ghrelin, and estradiol levels between EOC and control groups. EOC, epithelial ovarian cancer.

### Variable correlations

The correlation between estradiol, acylated, des-acylated and total ghrelin levels, on the one hand, and metabolic features of EOC-diagnosed women, on the other, is presented in Table 2. According to the results, estradiol levels and tumor size had a weak correlation with TG in the EOC group (*r* = 0.373 and *r* = 0.396, respectively). However, the results indicated that there was a negative significant correlation between estradiol and LDL-c as well as estradiol and HDL-c in the case group (*r* = -0.602, and *r* = -0.567, respectively). Interestingly, it seems that acylated ghrelin negatively correlated with estradiol levels (*r* = -0.226) among patients. However, no significant correlations were found between acylated, des-acylated, and total ghrelin, one the one hand, and measured metabolic features in EOC patients (*P* > 0.05 for all comparisons). Furthermore, acylated, des-acylated, total ghrelin and estradiol levels were found not to correlate with tumor stages or grades (*P* > 0.05 for all comparisons) as shown in Table 3.

**Table 2.** Correlation of estradiol, acylated ghrelin, des-acylated ghrelin and total ghrelin levels with metabolic features of women with EOC.

|  | Des-Acylated<br>ghrelin | Total ghrelin | Estradiol | Hemoglobin | Tumor Size | LDL-c | HDL-c | TG | TC |
| --- | --- | --- | --- | --- | --- | --- | --- | --- | --- |
| Acylated ghrelin | 0.2 | **0.2<sup>a</sup>** | **-0.2<sup>a</sup>** | 0.05 | -0.06 | -0.02 | 0.04 | 0.001 | -0.03 |
| Des-Acylated ghrelin |  | **0.9<sup>c</sup>** | 0.1 | -0.09 | -0.1 | -0.02 | -0.08 | -0.04 | -0.01 |
| Total ghrelin |  |  | 0.1 | -0.08 | -0.1 | -0.06 | -0.1 | -0.006 | 0.000 |
| Estradiol |  |  |  | 0.09 | **0.3<sup>a</sup>** | **-0.6<sup>c</sup>** | **-0.5<sup>c</sup>** | **0.4<sup>b</sup>** | 0.1 |

**a: Significant at 0.05, b: Significant at 0.01, c: Significant at 0.001**
EOC, epithelial ovarian cancer; TC, total cholesterol; TG, triglycerides; LDL-c, low-density lipoprotein cholesterol; HDL-c, high-density lipoprotein cholesterol.

**Table 3.** Comparison of estradiol, acylated ghrelin, des-acylated ghrelin and total ghrelin levels with tumor stages and grades in women with EOC.

|  | Stages |  |  |  | p<sup>a</sup> | Grades |  |  |  |  |  | p<sup>b</sup> |
| --- | --- | --- | --- | --- | --- | --- | --- | --- | --- | --- | --- | --- |
|  | Early (stage I-II) |  | Advanced (stage III–IV) |  |  | Grade I |  | Grade II |  | Grade III |  |  |
|  | Mean | SD | Mean | SD |  | Mean | SD | Mean | SD | Mean | SD |  |
| Acylated ghrelin | 7.4 | 2.3 | 9.8 | 5.6 | 0.3 | 10.4 | 5.4 | 7.5 | 3.2 | 8.5 | 3.3 | 0.1 |
| Des-Acylated ghrelin | 120.0 | 80.8 | 143.6 | 53.7 | 0.1 | 123.4 | 50.2 | 142.9 | 91.4 | 125.5 | 69.4 | 0.9 |
| Total ghrelin | 127.5 | 81.4 | 153.5 | 55.0 | 0.1 | 133.9 | 53.5 | 150.5 | 91.6 | 134.0 | 70.0 | 0.9 |
| Estradiol | 54.8 | 21.0 | 60.9 | 28.1 | 0.7 | 52.0 | 12.3 | 66.0 | 30.8 | 47.5 | 15.7 | 0.1 |
| P<sup>a</sup>: P-value Derived from Mann-Whitney Test. |  |  |  |  |  |  |  |  |  |  |  |  |
| P<sup>b</sup>: P-value Derived from Kruskal-Wallis Test. |  |  |  |  |  |  |  |  |  |  |  |  |

EOC, epithelial ovarian cancer.

### Logistic regression analysis

According to the logistic regression models, plasma acylated ghrelin was not significantly associated with the odds of EOC (OR: 0.96, 95% CI: 0.851-1.084, *P* = 0.512) in an age-adjusted model. Similarly, it was found that des-acylated and total ghrelin did not show a significant association with the EOC odds either (OR: 1.006, 95% CI: 0.996-1.017, *P* = 0.219 and OR: 1.007, 95% CI: 0.997-1.017, *P* = 0.165, respectively). However, estradiol level was reported to be significantly associated with the odds of EOC (OR: 1.083, 95% CI: 1.037-1.13, *P* < 0.001) as shown in Table 4.

**Table 4.**
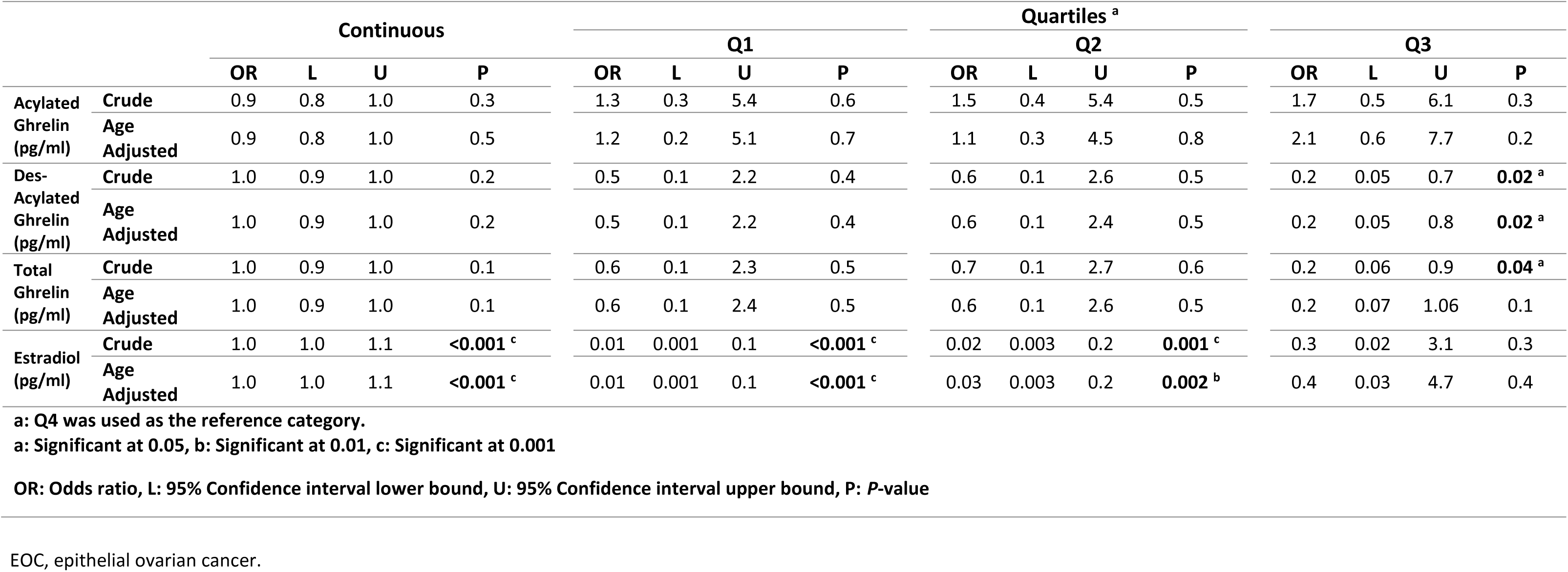
Association of plasma concentrations of acylated ghrelin, desacylated ghrelin, total ghrelin and estradiol and risk of EOC using logistic regression model.

The quartile analyses using age-adjusted model compared to the last quartile demonstrated that the des-acylated ghrelin levels in the third quartile (Q_3_) had a significant negative association with the EOC in both crude and age-adjusted models (OR: 0.192, 95% CI: 0.049-0.760, *P* = 0.019 and OR: 0.204, 95% CI: 0.051-0.812, *P* = 0.024, respectively). Nonetheless, total ghrelin levels were found to have a significant negative association with EOC odds only in the crude model (OR: 0.250, 95% CI: 0.066-0.950, *P* = 0.042). On the other hand, estradiol levels, in the two first quartiles (Q_1_, Q_2_), indicated an negatively significant association with the EOC odds in both crude and age-adjusted models (OR: 0.010, 95% CI: 0.001-0.104, *P* < 0.001 and OR: 0.011, 95% CI: 0.001-0.118, *P* < 0.001 for the first quartile and OR: 0.024, 95% CI: 0.003-0.226, *P* = 0.001 and OR: 0.030, 95% CI: 0.003-0.284, *P* = 0.002 for the second quartile, respectively).

## DISCUSSION

OC is the most lethal malignancy among women, and the involvement of estrogen in OC development and progression is examined by many researchers (20). On the other hand, various studies have shown ghrelin’s effect on cancer development through cell proliferation, tissue invasion, and metastasis (21). Thus, the present study investigated estradiol, acylated, des-acylated and total ghrelin levels in postmenopausal patients with EOC. The main finding of the present study is that estradiol plasma levels were related to a significant increase in postmenopausal patients with EOC, implying that higher levels of estradiol are associated with EOC occurrence. The current findings suggest that each unit of increase in estradiol levels will raise the EOC odds up to about eight percent in postmenopausal women. However, interestingly, higher levels of estradiol in the first two quartiles compared to the last quartile showed a protective effect on the EOC odds, indicating that lower estradiol levels may have a preventive effect on EOC in these women. Furthermore, regression analysis indicated that higher levels of des-acylated and total ghrelin levels in the third quartile decrease the EOC odds. An ample body of studies has shown that estradiol, a subgroup of estrogen, plays important roles in OC incidence (9, 22, 23). Estrogen plays its part not only through interaction with intracellular receptors, which modulate gene expression, but it also acts via its membrane receptor and cytoplasmic signaling cascades related to cell growth, maintenance, and differentiation (24). The current findings showed significantly higher levels of estradiol in EOC patients in comparison with normal participants. Moreover, similar results were obtained from previous studies regarding the levels of estradiol in breast and OCs (25). Furthermore, recent studies reported the role of estradiol in cancer progression and invasion through several different mechanisms, including cell proliferation, involvement in apoptosis procedure, angiogenesis, and migration (26, 27). Thus, all these findings support estradiol’s role in development and progression of OC. In contrast, other evidence suggested that estrogens including estradiol may not be the most relevant etiologic factor for OC incidence. It is believed that an estrogen-raising procedure like pregnancy not only does not increase the OC risk but also has a protective effect, therefore, some other etiologic factors must be involved in OC (28). The current results obtained from the regression analysis also demonstrated that despite estradiol’s role in increasing the EOC odds at high levels, lower estradiol levels showed a protective effect on EOC in postmenopausal women, as some studies indicated beneficial effects of low-dose oral contraceptives consumption, including low dosages of ethinyl estradiol (29). Thus, in spite of all detrimental effects of estrogen presence on cancer progression, it is suggested that expression of a known promoter of angiogenesis, angiopoietin-1, is reduced by estradiol in ER-α^+^ breast cancer (30). As presented in the previous studies, the cellular response to estradiol stimulation does not always cause cancer progression and in some cases may be a beneficent factor such that estradiol can block ER-α^+^ expression through specific mechanisms (26, 29).

The Pearson and Spearman correlation analyses revealed the positive correlation between estradiol and des-acylated ghrelin levels among EOC patients, thereby suggesting the protective effect of higher des-acylated ghrelin levels on postmenopausal patients with EOC due to decrease in estradiol levels. Moreover, estradiol levels positively correlated with ovarian tumor size in the present study. To our knowledge, this correlation has not been investigated by previous studies thus far. Meanwhile, no correlation was indicated between estradiol levels and cancer stages or grades in the present study, while conflict results indicated a positive association between estradiol levels, tumor volume, and cancer stage (31, 32). The present study also revealed a negative correlation between estradiol and patient’s LDL-c and HDL-c; however, there was a positive association between estradiol and TG. In contrast to the related literature, a study conducted by Bojar et al. reported a negative correlation between estradiol and LDL-c levels. Nonetheless, estradiol and HDL-c were reported to be positively correlated among OC patients (33).

Ghrelin, as an adipocyte-derived hormone, is expressed in a wide range of cancer tissues and plays a pivotal role in a number of key procedures in cancer progression, including cell proliferation, cell migration and invasion, and apoptosis (15). Of few studies performed in this field, Markowska et al. showed that blood concentrations of acylated ghrelin were higher in OC patients. In contrast, total ghrelin blood concentrations showed no significant difference among the studied groups (34). The findings obtained in the present study do not coincide with those of the above-mentioned studies as in this study. It was indicated that high levels of total and des-acylated ghrelin indicated a protective effect on EOC in postmenopausal women; however, no correlation was observed between acylated, des-acylated and total ghrelin, on the one hand, and tumor size, cancer grade, and stage, on the other. While previous studies, which investigated the association of ghrelin levels and histopathological factors such as tumor grade and differentiation, indicated a positive correlation between total serum ghrelin, tumor grade and differentiation in colon cancer, there was a negative correlation between ghrelin levels and histologic grades in endometrium carcinomas (35, 36). An explanation of the contradictory findings of the present study and those of other ones may be the fact that in this study plasma levels of acylated, des-acylated, and total ghrelin were measured, whereas most of the other studies measured total ghrelin. These novel findings suggest that ghrelin may have an important role in EOC prevention through several mechanisms. The biological actions of ghrelin have been studied in the male gonad, which shows ghrelin’s effect on the modulation of steroidogenesis and, probably, the proliferation of interstitial Leydig cells; however, the functional role of ghrelin in the ovary has not been explored well (37, 38). Despite all ghrelin’s peripheral actions, several studies have suggested that ghrelin plays a role in controlling cell proliferation and cancer (39). However, there are numerous conflicting reports, which suggest a ghrelin-mediated inhibiting role in the growth of cancer cell, including the OC cells known as HO-8910 as well as thyroid, lung, and breast cancer cell lines (16, 40, 41). This protective effect of ghrelin and des-acylated ghrelin may be mediated through ERK1/2 branch of MAPK pathways (41, 42). Studies also show that des-acylated ghrelin had a more pronounced protective effect than acylated ghrelin, which confirmed the present results (43).

To the best of authors’ knowledge, this is the first attempt made to examine the levels of plasma estradiol, acylated, des-acylated and total ghrelin in postmenopausal patients with EOC. The minor limitations of the present study include the relatively small sample size and its single-center design. Since a case-control design cannot distinguish between biomarkers that increase odds of developing the disease and biomarkers levels that are caused by disease onset, further cohort and prospective studies are highly recommended.

## Conclusions

In summary, the present study provides evidence that postmenopausal women suffering from EOC may show higher estradiol levels than their counterparts. The authors suggest that high levels of estradiol might be a potential odds factor in EOC, and that lower estradiol levels have a protective effect on EOC development. Notably, it is proposed that higher levels of des-acylated and total ghrelin may protect patients against EOC. The current findings underscore the need for further investigations to explore other probable etiologic factors involved in estradiol and ghrelin’s odds/protective effects on EOC.

## Data Availability

The dataset of the current study are available from the corresponding author on reasonable request.

## Acknowledgement

Authors would like to express their deep gratitude towards participants who provided us with their precious assistance in performing this study.

